# Prevalence of ABO and Rhesus blood groups in Blood donors: A retrospective study from a tertiary care blood bank in Pondicherry

**DOI:** 10.1101/2024.05.14.24306971

**Authors:** Kingsley Simon, Jain S Rahul, S Manjineesha

## Abstract

**Introduction:** The ABO blood group system was the first human blood group system discovered by Austrian scientist Karl Landsteiner in 1901. The ABO blood group system is divided into four blood types on the basis of presence or absence of A and B surface antigens A and B antibodies presence in the plasma. Positive and Negative of the blood group determines by presence or absence of the D antigen..

**Objectives:** This study was carried out to determine the distribution pattern of blood groups among blood donors at a tertiary care hospital of Puducherry

**Materials and Methods:** It is a retrospective study done a period of 5 years from May 2017 to May 2022 that includes the 9062 blood donors. Blood group done as per the standard operative procedure. Data on the ABO and Rh blood group type of all blood donors was collected and analysed.

**Result:** The most common ABO blood group type was Group O (38%) followed by group B (33.5%), group A (21.6%) and group AB (6.9%) and one Bombay blood group. The distribution of Rh(D) blood group type revealed 92.5% as Rh-positive and 7.5% as Rh-negative.

**Conclusion:** The most common ABO blood group type in our region (Puducherry) is Group O followed by B and A while, AB and Bombay groups are the least common type. Knowledge of the blood group distribution pattern is essential for the effective delivery of blood banking services.

## INTRODUCTION

Blood group system antigens are present on the red cell membrane. Antigens are inherited from our parents in Mandelian fashion and express on the RBC membrane early in life and remain same throughout the life. ABO antibodies are naturally occurring (IgM) antibodies present in the plasma. In 1900 Austrian scientist Karl Landsteiner originally described the ABO blood group system (1). In 1902 Von Decastello and Sturli discovered the AB blood group(2). ABO genes located on long arm of chromosome 9, these genes produces galactosyl transferase enzymes. With the help of enzymes blood group antigens are expressed on the surface of red cell membrane and many tissues such as heart, lung, kidney and endothelium. A and B antibodies develop after 3-6 months of age and it is not present at birth. (1).

ABO and Rh blood group generally detected by presence or absence of antigens present on the red cell membrane and absence of the corresponding antibodies in the plasma.

According to the blood group antigens and antibodies ABO system divided into four blood group types, such as A, B, O and AB.

More than 50 Rh antigens have been discovered in the Rh system. Five principle antigens are clinically significant which includes D, C,c,E, and e. Among the Rh antigens, D antigen is more immunogenic. It is extremely crucial in obstetrics and being the main cause of haemolytic disease of the newborns (HDN). Positive and negative group refers to presence or absence of a D antigens (2).

Rh antibodies are IgG type antibodies and developed after the sensitizing events, such as transfusion, transplantation and pregnancy.

The ABO and Rh blood groups are important for blood transfusion practice, but they are also helpful in clinical research, population genetic studies, population migration patterns study, and settling some medico legal disputes, especially those involving cases of disputed paternity. (3).

For the effective management of blood bank inventory, we need to know the prevalence of f ABO and Rh blood group system in our community. The prevalence of the ABO blood group system is significantly influenced by race, ethnicity, and geographic location (4).

Because it is crucial to understand how blood types are distributed in our population, this study has been conducted to observe the prevalence of the ABO and Rhesus blood types distributed among our blood donors.

## MATERIALS AND METHOD

It is a retrospective study that was conducted at PIMS Blood Center in Pondicherry from May 2017 to May 2022. According to the guideline established by the Drugs and Cosmetics Act 1940 and rules 1945, a total of 9062 donors were selected. This included donors who had volunteered to donate blood as well as those donating for their patients as a replacement.

### Ethics and Informed Consent

Our study was approved by the ethics committee of PIMS INSTITUTE ETHICS COMMITTEE (IEC). REF NO: IEC: RC/18/107

### Selection of Donors

Donors selected according to GSR 166E of Drugs and Cosmetic acts 1940.

### Blood grouping of the donors

Forward and reverse grouping done by the conventional tube method according to blood bank’s standard operative procedures. Forward grouping was done with monoclonal anti A, anti B, anti H antisera (Tulip diagnostics ltd) and reverse grouping was done with in-house prepared A, B, O pooled cells.

All the negative group donors proceeded to Du testing to confirm the Rh D antigen. If the Du testing is positive, donor considered to be positive blood group.

The forward grouping was done by one technologist and reverse grouping by another technologist. The blood type was verified only if reverse grouping showed the same result as forward grouping. All the reagents were utilised only after performed the quality control according to drug and cosmetic act rules 1945.

Donor blood group was collected from blood group donor register and blood donor details (Age, sex, voluntary and replacement) were collected from blood donor register.

## STATISTICAL ANALYSIS

All the data was entered into Microsoft Office 2010’s Excel sheet. For the analysis descriptive statistics like mean and percentages were calculated for the analysis.

## RESULTS

Total number of donors were 9062. Of which male donors were 8919(98.4%) and the female donors 143(1.6%). Voluntary donors were 6664 (73.5%) and replacement donors were 2398(26.5%).

Mean age of the donor was 27.6 years. The majority of donors were in the 18- to 25-years 4225 (46.6%) (Table 1).

**Table 1:**
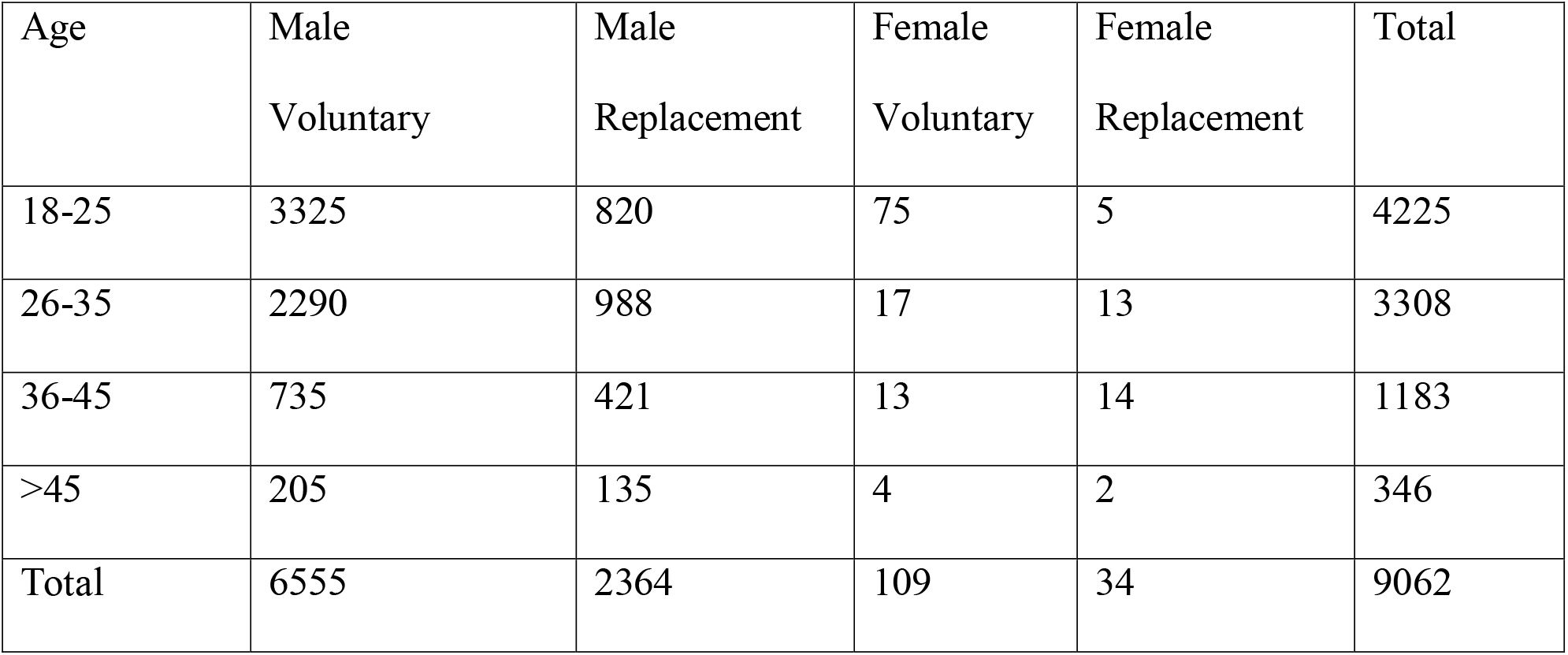
Distribution of age, gender and type donors.

| Age | Male<br>Voluntary | Male<br>Replacement | Female<br>Voluntary | Female<br>Replacement | Total |
| --- | --- | --- | --- | --- | --- |
| 18-25 | 3325 | 820 | 75 | 5 | 4225 |
| 26-35 | 2290 | 988 | 17 | 13 | 3308 |
| 36-45 | 735 | 421 | 13 | 14 | 1183 |
| >45 | 205 | 135 | 4 | 2 | 346 |
| Total | 6555 | 2364 | 109 | 34 | 9062 |

The most predominant blood group Among the 9062 blood donors was “O” (38%) followed by “B” (33.5%) then “A”(21.6%), “AB” (6.9%) and one Bombay blood group donor. In the Rh D factor, predominant donors were Rh positive were 8386 (92.5) and Rh negative were 676 (7.5) (Table 2).

**Table 2:**
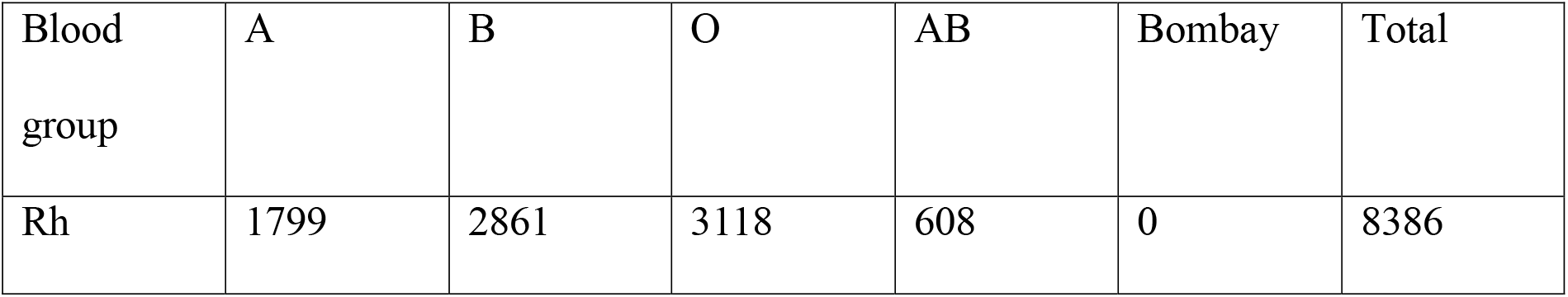
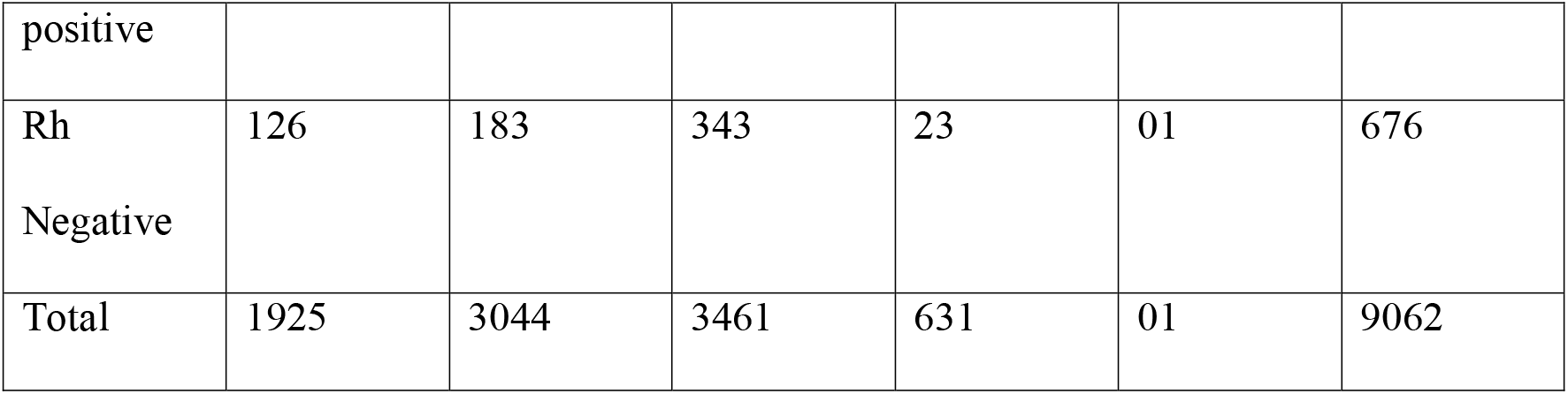
Frequency of blood grouping and Typing.

Among the total deferred female donors, 78 (83.8%) deferred due to low haemoglobin levels (Table 3).

**Table 3:** Donor deferral.

| Reasons | Males | Females | Total |
| --- | --- | --- | --- |
| Low haemoglobin | 41 | 78 | 119 |
| Others | 347 | 15 | 362 |
|  | 388 | 93 | 481 |

Comparison of blood grouping and typing of our study results with other studies shown in table4.

**Table 4:**
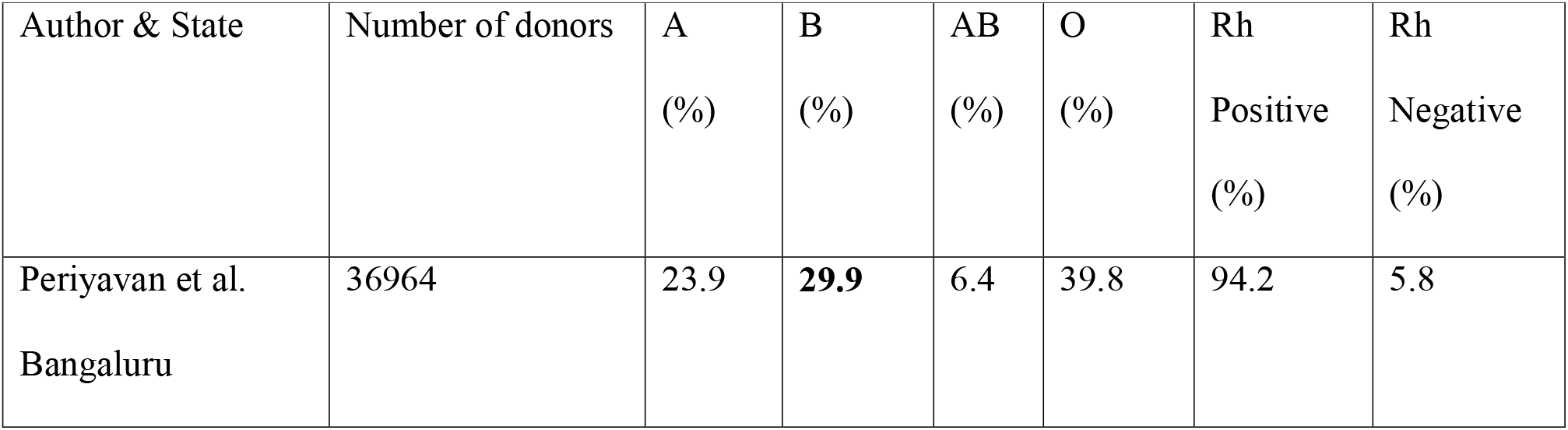
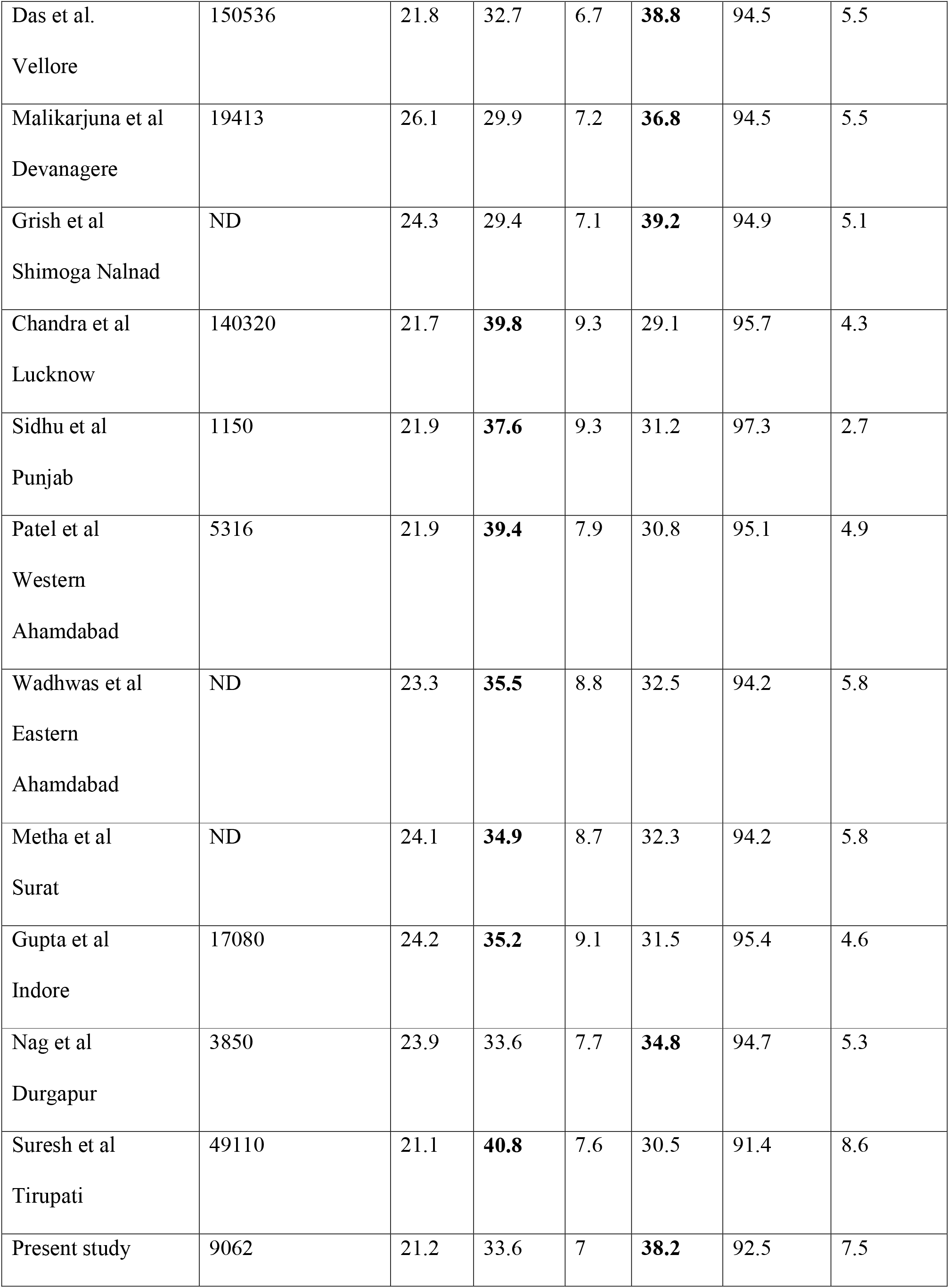
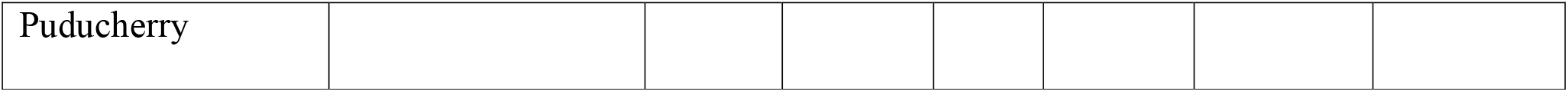
Distribution (%) of ABO & Rh phenotypes at different regions India.

During the study period, 15160 blood components were distributed, with blood group “O” and blood group “B” being the most common.(Table 5).

**Table 5:** Blood components issued with blood group.

| Blood group | A | B | O | AB | Bombay | Total |
| --- | --- | --- | --- | --- | --- | --- |
| Rh positive | 2965 | 4756 | 5315 | 1108 | 0 | 14144 |
| Rh Negative | 176 | 305 | 466 | 68 | 01 | 1016 |
| Total | 3141 | 5061 | **5781** | 1176 | 01 | 15160 |

## DISCUSSION

In our study male donors (98.4%) contributions was more than the female donors (1.6%) which is similar with other Indian studies (5, 6, 7). This is generally due to societal stigma, cultural customs and lack of drive and fear of giving blood, among the females in the community. In our study, among the female deferred donors, 83.8% deferred due to low haemoglobin. Thus, a healthy nutritious diet and iron supplements are necessary to improve the overall health of females. Female donors need to be inspired and made aware of the benefits while also being freed from misconceptions and anxieties.

Most of the blood donors under the age group of 18-25 years. This is comparable with other studies (5,8). The age group above 45 years had the fewest donors since so many persons in this age range have chronic health problems, low haemoglobin, hypertension, and diabetes mellitus, which prevent them from giving blood or cause them to be deemed unfit during predonation counseling.

In our country the number of voluntary blood donors increased from 54.4% in 2006–2007 to 83.1% in 2011–2012 (9). Parul Garg *et al* from Uttarkhand, observed in their study, 99.7% were replacement donors (10). In our study voluntary donors were 73.5%. It is necessary to increase the number of voluntary donations so that all patients in need of blood can receive it immediately without having to wait for replacement donors. Moreover, voluntary donations reduce the risk of transfusion-transmitted diseases. (11). The voluntary contributions must be encouraged in order to obtain safe blood.

According to the current study, the blood type “O” (38%) is the most prevalent, followed by “B” (33.5%), “A” (21.6%), and “AB” (6.9%). Contrary to earlier research from North India, where it was shown that blood group B predominated, this was incongruous. (11,12,13) Studies by Periyavan *et al*. and Suresh *et al*. from South India showed a comparable frequency trend where the O blood group predominated. (14,15).

The most common blood types in the neighbouring nations like Pakistan and Nepal were B and A, respectively (16,17).

In a large, multicentric survey, blood group “O” donors made up the majority (37.1%), followed by blood groups B (32.3%), A (22.9%), and AB (7.7%), which was the least common. The results were consistent with our study and those anticipated by the Asiatic trend, which shows that O is the most common letter followed by B, A, and AB (14)

The prevalence of Bombay blood group in our study was found to be 0.011%. This is little higher than the Tamil Nadu (0.004%) and Karnataka (0.005%) studies and lesser than the study from Suresh *et al* (0.03%) (15). In a study from Talukder B *et al* (18). Bombay phenotypic prevalence was observed to be 0.011% overall, with a prevalence of 0.007% in donors and 0.014% in patients, respectively. About 1 in 10,000 Indians and 1 in 100,000 Europeans, respectively, have the Bombay blood group in the general population. (19) The Kutia Kondh tribe in Orissa, eastern India, has a high incidence of the Bombay phenotype (20). One in 278 people in the Bhuyan tribal population, according to another study (21) from northwest Orissa, have the Bombay phenotype. Bombay blood group detection, which is sometimes misconstrued for the “O” group, requires population surveys. All blood banks must keep a thorough register of these uncommon blood groups so that they can be used in case of emergencies.

In much of India, the prevalence of the Rh D blood group ranges from 94% to 98%, while that of the Rh D negative blood group ranges from 2% to 6%. (22). Rh-negative phenotypes affect 15% to 17% of White people, but are less prevalent in other ethnic groups (23). In the current study, 92.5% of the donors were Rh D positive, while 7.5% were Rh D negative.

We analysed the blood components distributed in accordance with blood group. It was discovered that “O” group blood components were primarily used. This is one of the reason O group is predominant in our study.

Understanding blood group systems makes it easier to manage the local blood bank and transfusion services in case of emergencies. The researchers could perhaps investigate the factors influencing the observed blood group system distribution patterns using the data obtained.

## CONCLUSIONS

This study shows that among blood donors, blood group “O” is the most prevalent and blood group “AB” is the least prevalent. Rh positive donors made up 92.5% of the Rhesus blood group system, while Rh negative donors made up 7.5%. Female blood donors were extremely low, and it needs to be raised by raising health awareness about blood donation among women. Health planners can prepare for the region’s future health concerns by using the data from this study and several other studies of various geographical regions of India.Every individual’s blood group must be listed on their national identification card, driver’s licence, and identity card of school or the workplace. This will be extremely helpful in situations where an urgent transfusion of blood that is yet to be cross-matched is necessary. Similar, well-designed studies must be carried out in other Indian states to ascertain the blood group frequencies there.

## Data Availability

All data produced in the present study are available upon reasonable request to the authors

